# The impact of SARS-CoV-2 variants on the likelihood of children identified as sources of infection in the NIH workforce: a cohort study

**DOI:** 10.1101/2023.11.07.23297422

**Authors:** Jessica M. van Loben Sels, Heike B. Bailin, Michael R. Bell, Jessica McCormick-Ell, Sanchita Das, Allison E. Roder, Elodie Ghedin, Michael McGann, Amanda D. Castel, D. Rebecca Prevots, Jennifer L. Kwan

**Author notes:** Corresponding author at: 5601 Fishers Lane, Epidemiology Unit, Laboratory of Clinical Infectious Disease, NIH, Rockville, MD, USA.

## Abstract

**Background:** Children (<18 years old) were not initially considered significant sources of infection (SOIs) for SARS-CoV-2. Risk mitigation strategies were thus prioritized for adults, and vaccination was inaccessible for children until mid-2021. Emergence of novel variants led to significant increases in COVID-19 cases in both children and adults. Whether these emergence events and increased vulnerability of unvaccinated children had a synergistic effect resulting in increased caseloads in adults requires further exploration.

**Methods:** A retrospective cohort study was conducted among 3,545 workers diagnosed with COVID-19. Case details were compiled during contact investigations. Variants of concern were identified following sequencing of biological samples collected through employer-based testing programs. Logistic regression was performed to compare the odds of having a child SOI based on the dominant variant in the workforce.

**Results:** One-fourth (24.5%) of the cohort reported having a child in-residence; 11.2% identified a child as their SOI. In Alpha-dominant months, the odds of having a child SOI were 0.3, and the child SOI was likely older (5-17 years old). The odds of having a child SOI increased to 1.3 and 2.2 in Delta- and Omicron-dominant months, respectively. The odds of having younger child SOIs (<5 years old) were significantly higher in Omicron-dominant months.

**Conclusions:** Children were highly likely to acquire the virus and posed a significant risk of transmission to their adult caretakers during Delta- and Omicron-dominant months. Without proper mitigation strategies in both the home and the workplace, child-associated transmission can threaten operations in the forms of staff shortages.

**What is already known on this topic:** Increases in transmission trends related to SARs-CoV-2 Variants of Concern have been documented in the literature at the population level and in workplaces.

**What this study adds:** This study looks more closely at the role that children played in transmission to adult workers, and therefore their potential to seed transmission outside of the home. This interface of transmission has been neglected in the literature but is key for future policy development.

**How this study might affect research, practice, or policy:** Transmission of SARS-CoV-2 from children to their caretakers may cause significantly increased odds of infection in a worker population. This may have second order effects for staffing, particularly in workgroups with employees of childbearing age. Employers should consider this in the design of their policies for continuity of operations, telework, and leave.

## Background

Early in the pandemic, children under the age of 18 were generally considered low risk sources of infection (SOIs) for SARS-CoV-2 (1). Their period of illness was typically limited with rare severe cases warranting hospitalization (2, 3). Therefore, older individuals were prioritized for COVID-19 vaccination (4, 5) in an age hierarchy that reflected perceived risk of severe disease, transmission potential, and clinical trial timelines (6, 7) (Figure 1A).

**Figure 1.**
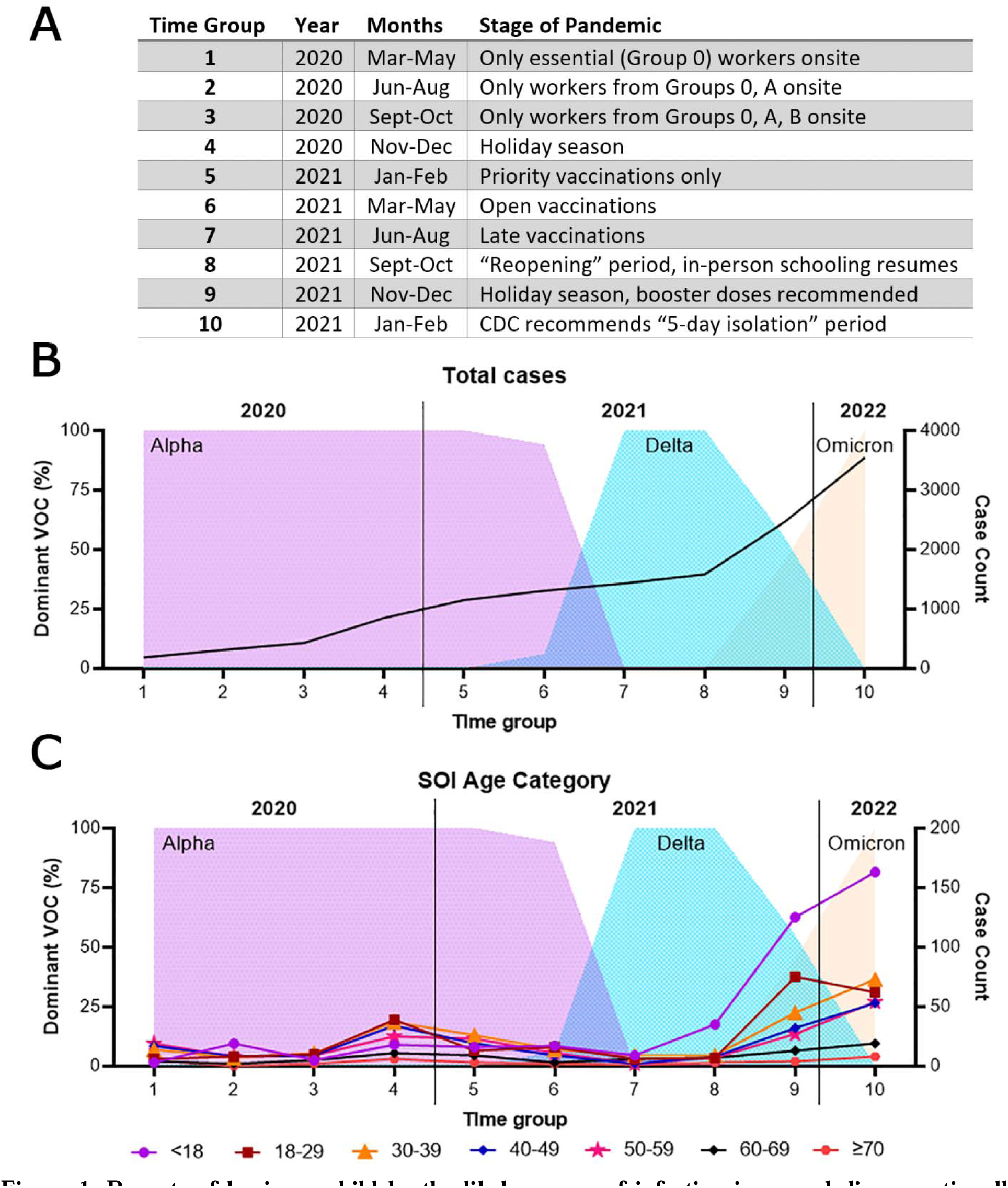
Reports of having a child be the likely source of infection increased disproportionally around the emergence of the Delta variant in the study population. A) Rationale used to group months together into time groups based on changes employed at the NIH or within the community that may affect case counts. B) Variants of concern detected within the NIH workforce, represented in time groups and as percentages of total samples sequenced, overlayed with cumulative NIH worker case counts. C) Variants of concern detected within the NIH workforce, represented in time groups and as percentages of total samples sequenced, overlayed with cumulative case counts stratified by SOI age categories.

In 2020 and the first 6 months of 2021, COVID-19 cases worldwide were predominantly caused by ancestral SARS-CoV-2 strains that later became dominated by the B.1.1.7 Alpha variant (Figure 1B) (8). This variant of concern (VOC) was displaced by the Delta variant in summer 2021 (9) and coincided with increases in adult vaccination (10), whereas children lagged behind. In the last months of 2021, the Omicron variant was dominant (11). Omicron continues to be the dominant VOC as of early-2023 and has resulted in predominantly symptomatic infections in all ages independent of vaccination status, though severity of illness was significantly decreased among vaccinees (12).

The National Institutes of Health (NIH) is a federal agency whose workforce was impacted by the SARS-CoV-2 pandemic and benefitted from onsite testing programs and in-depth, workplace contact tracing. All workers newly diagnosed with COVID-19 were assessed by the agency-based occupational medical service (OMS) to supplement local health departments and characterize workplace risk factors. One such risk was the workers’ suspected source of infection (SOI). While SOIs could not be clearly established in all cases, 65% of workers with COVID-19 could identify a close contact with symptoms or a positive test predating their own positive test and/or symptom onset. The likely SOI’s age and relationship to the worker were recorded during contact tracing interviews.

This study aimed to determine if increases in child SOI assignment by age (Figure 1C) were significant and if the emergence of new dominant VOCs contributed to increased transmission risk to staff. Such findings would refute the early assumptions that children were not at a high risk of contracting COVID-19 and therefore were not significant reservoirs for transmission.

## Methods

### Study Population

The target population of this study was adult caretakers of children, and the study population was the NIH workforce comprised of nearly 50,000 people with worksites in several locations across the United States (Supplementary Table 1). From March 2020 through February 2022, more than 4,500 workers with positive COVID-19 tests (i.e., cases) were reported. This retrospective cohort study included NIH workers who reported a positive COVID-19 test to OMS or tested positive through a workplace testing program. Those excluded were (1) positive cases originating from the NCI Frederick, MD, site which were investigated separately, and (2) patients at the NIH Clinical Center and their visitors (non-staff). After these exclusions, our study population comprised 3,545 NIH workers, a likely an underestimation of the total burden experienced by the NIH workforce due to the voluntary reporting of positive results from community-based or at-home testing.

### Testing and laboratory procedures

The OMS received positive results through three testing mechanisms, as outlined in Figure 2. Workers were instructed to report any positive COVID-19 test conducted in the community through an online survey. Testing was also offered to staff via two programs: “for-cause” or asymptomatic based on risk assessment. The “for-cause” testing prioritized staff with recent symptom onset or high-risk exposure and sampled via a drive-through testing line. Nasopharyngeal or mid-turbinate nasal swabs were collected by qualified staff and prioritized for analysis by the NIH Clinical Center (CC) Department of Laboratory Medicine (DLM). This NIH-based mode of testing was offered only on the main campus in Bethesda, MD. Conversely, all NIH sites had asymptomatic, surveillance-based testing programs established through which either mid-turbinate nasal swabs were collected by trained staff or saliva samples were self-collected. Healthcare personnel were strongly encouraged to test once a week until fully vaccinated. For all other workers, testing was at their discretion.

**Figure 2.**
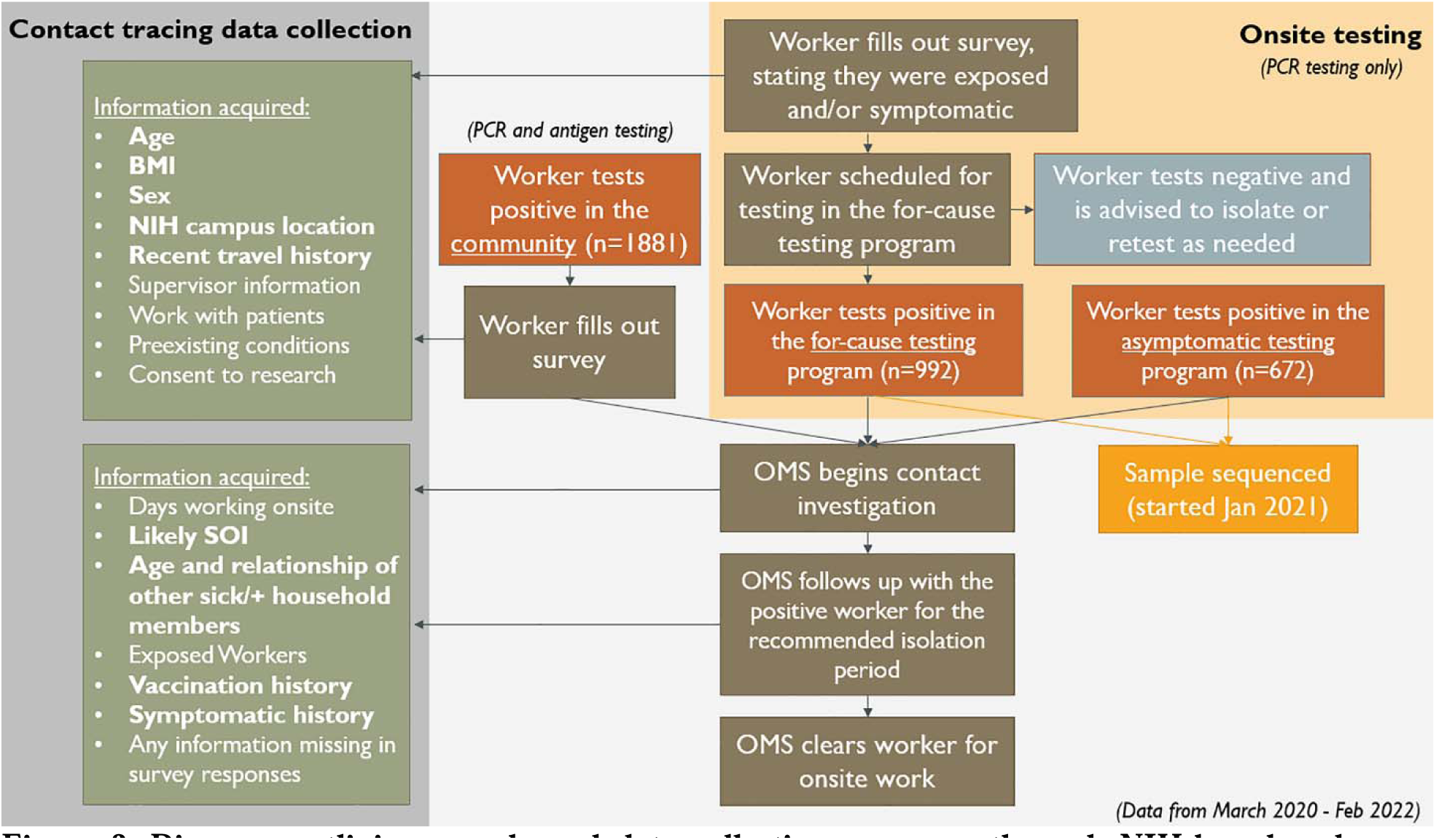
Diagram outlining sample and data collection processes through NIH-based and non-NIH-based, community testing programs.

All NIH-based COVID-19 testing followed established protocols (13). Samples collected from the “for-cause” testing program were assayed individually by PCR, while samples collected from surveillance programs were pooled and deconvoluted if the pool tested positive for the SARS-CoV-2 RNA via PCR. Any result with a cycle threshold (Ct) value below 40 (a measure of viral load) was reported as a positive result to the OMS. For samples with Ct values under 30, RNA was extracted from the sample, reverse transcribed, and PCR-amplified using a modified version of the ARTIC protocol (14) which includes primers spanning the whole SARS-CoV-2 genome (15). Amplicons were sequenced via Illumina, and reads were assembled into a genome then compared to published genomes using Pangolin (16). All assembled genomes were submitted to GISAID. VOC designation followed the WHO naming system (17). Samples with Ct values higher than 30 could not be sequenced due to low viral loads.

### Data collection and statistical analysis

Analysis was conducted using deidentified data that were collected for public health purposes thus exempted from IRB. Original data collection required consent of the employee in accordance with the OMS Privacy Act Notice and System of Records Notice which explained that samples could be used in research protocols in a deidentified manner.

As diagramed in Figure 2 and outlined in Supplementary Table 1, data were collected via self-reported electronic survey which requested demographic information (e.g., sex, weight, height, vaccination status, etc.) and reason for submitting the request: whether they suspected exposure, tested positive elsewhere or had symptoms. Staff reporting positive results were followed up by OMS-administered contact tracing interviews completed by telephone, collecting information: suspected source of infection, travel history, onsite presence, and other exposures. Medical histories and workplace COVID-19 testing records supplemented these data.

Each case was followed for the duration of their symptoms and until they met clearance criteria for returning to work, per evolving CDC recommendations. The following variables were extracted from open text fields in contact tracing reports: age and relationship of household members, suspected SOI, suspected SOI relationship, and travel history. Remaining variables were categorized by the contact tracer during interview.

Descriptive statistics were generated for unstratified data and data stratified by having a child in-residence (CIR) and having a child SOI. Continuous variables were categorized prior to analyses, and categorical variables were reported with frequencies and percentages. Logistic regression was used to assess the relationships of dominant VOCs to CIR and to child SOI by determining odds ratios (ORs), 95% confidence intervals (CIs), and significance (p <0.05). Time-dependent variables were addressed in final model adjustment to avoid redundancy. All statistical analyses were conducted using SAS version 9.4.

## Results

### Characteristics of study population

From March 2020 to the end of February 2022, 3,545 cases were identified and met the criteria of our study (Table 1). One fourth of positive employees reported having a child in-residence (CIR, 25.1%), and less than half of CIR cases assigned the child as the likely SOI (11.2% of total cases, 41.8% of CIR cases). A higher percentage of men reported having a CIR (56.9%), but a higher percentage of women reported a child was their likely SOI (63.0%). Most CIR and child SOI cases were reported by workers aged 30 through 50 years (69.3% and 73.5%, respectively). About 67% of both CIR and child SOI cases were onsite during their POIs. Cases with CIR and child SOI cases were less likely to test positive through asymptomatic testing (15.8% and 11.3%, respectively), and more CIR and child SOI cases were vaccinated at the time they tested positive (66.8% and 77.6%, respectively, versus 59.1% of cases overall). However, vaccination patterns changed significantly over time and were accounted for in our modeling.

**Table 1.** Descriptive statistics for the study population, stratified by outcomes of interest. Data were generated three ways: (1) unstratified, (2) stratified by whether a child was living in the same household as the positive case (CIR), and (3) stratified by whether a child was determined to be the likely source of infection (SOI) for the positive NIH worker. Counts and percentages are reported for each stratum.

| Variable | Cumulative | Child in-residence (CIR) |  | Child SOI |  |
| --- | --- | --- | --- | --- | --- |
|  |  | Yes | No | Yes | No |
| **Total cases** | **3545 (100)** | **890 (25.1)** | **2675 (74.9)** | **400 (11.2)** | **3145 (88.8)** |
| Sex |  |  |  |  |  |
| Male | 1722 (48.6) | 495 (56.9) | 1347 (50.4) | 148 (37.0) | 1574 (50.0) |
| Female | 1823 (51.4) | 375 (43.1) | 1328 (49.6) | 252 (63.0) | 1571 (50.0) |
| Age of IC |  |  |  |  |  |
| 18-29 | 710 (20.0) | 54 (6.2) | 656 (24.5) | 26 (6.5) | 684 (21.8) |
| 30-39 | 1015 (28.6) | 303 (34.8) | 712 (26.6) | 138 (34.5) | 877 (27.9) |
| 40-49 | 761 (21.5) | 300 (34.5) | 461 (17.2) | 156 (39.0) | 605 (19.2) |
| 50-59 | 648 (18.3) | 164 (18.9) | 484 (18.1) | 64 (16.0) | 584 (18.6) |
| 60-69 | 346 (10.0) | 43 (4.9) | 303 (11.3) | 14 (3.5) | 332 (10.6) |
| ≥70 | 52 (1.5) | 6 (0.7) | 46 (1.7) | 2 (0.5) | 50 (1.6) |
| Obese? (≥30 BMI) |  |  |  |  |  |
| Yes | 710 (20.0) | 201 (23.1) | 509 (19.0) | 101 (25.2) | 609 (19.4) |
| No | 1646 (46.4) | 394 (45.3) | 1252 (46.8) | 179 (44.8) | 1467 (46.6) |
| No info | 1189 (33.5) | 275 (31.6) | 914 (34.2) | 120 (30.0) | 1069 (34.0) |
| NIH site |  |  |  |  |  |
| Main campus (MD) | 2458 (69.3) | 608 (69.9) | 1850 (69.2) | 288 (72.0) | 2170 (69.0) |
| Satellite sites (MD) | 825 (23.3) | 205 (23.6) | 620 (23.2) | 84 (21.0) | 741 (23.6) |
| NIEHS (NC) | 138 (3.9) | 24 (2.8) | 114 (4.3) | 6 (1.5) | 132 (4.2) |
| RML (MT) | 106 (3.0) | 23 (2.6) | 83 (3.1) | 17 (4.3) | 89 (2.8) |
| NIDDK (AZ) | 18 (0.5) | 10 (1.2) | 8 (0.3) | 5 (1.2) | 13 (0.4) |
| Hire Type |  |  |  |  |  |
| Fellow | 571 (16.1) | 63 (7.2) | 508 (19.0) | 38 (9.5) | 533 (17.0) |
| Employee | 1477 (41.7) | 472 (54.3) | 1005 (37.6) | 224 (56.0) | 1253 (39.8) |
| Contractor | 1415 (39.9) | 320 (36.8) | 1095 (40.9) | 132 (33.0) | 1283 (40.8) |
| Other | 82 (2.3) | 15 (1.7) | 67 (2.5) | 6 (1.5) | 76 (2.4) |
| Work Type (RTWG) |  |  |  |  |  |
| Administration (C) | 1043 (29.4) | 295 (33.9) | 748 (28.0) | 116 (29.0) | 927 (29.5) |
| Animal care (0) | 295 (8.3) | 54 (6.2) | 241 (9.0) | 23 (5.8) | 272 (8.7) |
| Clinical (0) | 441 (12.4) | 158 (18.2) | 283 (10.6) | 89 (22.3) | 352 (11.2) |
| Housekeeping (0) | 102 (2.9) | 26 (3.0) | 76 (2.8) | 7 (1.8) | 95 (3.0) |
| Maintenance (0) | 356 (10.0) | 76 (8.7) | 280 (10.5) | 36 (9.0) | 320 (10.2) |
| Research (A/B) | 1084 (30.6) | 208 (23.9) | 876 (32.8) | 109 (27.3) | 975 (31.0) |
| Security (0) | 156 (4.4) | 37 (4.3) | 119 (4.5) | 15 (3.8) | 141 (4.5) |
| Onsite during POI? |  |  |  |  |  |
| Yes | 2541 (71.7) | 587 (67.5) | 1954 (73.1) | 267 (66.8) | 2274 (72.3) |
| No | 1004 (28.3) | 283 (32.5) | 721 (26.9) | 133 (33.2) | 871 (27.7) |
| Source of infection |  |  |  |  |  |
| Community | 1242 (35.0) | 170 (19.5) | 1072 (40.1) | 3 (0.8) | 1239 (39.4) |
| Household | 1313 (37.0) | 551 (63.3) | 762 (28.5) | 369 (92.3) | 944 (30.0) |
| Non-household | 400 (11.3) | 60 (6.9) | 340 (12.7) | 28 (7.0) | 372 (11.8) |
| Travel | 308 (8.7) | 51 (5.9) | 257 (9.6) | 0 (0.0) | 308 (9.8) |
| SOI relationship |  |  |  |  |  |
| Significant other | 367 (10.4) | 91 (10.5) | 276 (10.3) | 0 (0.0) | 367 (11.7) |
| Parent | 106 (3.0) | 17 (2.0) | 89 (3.3) | 0 (0.0) | 106 (3.4) |
| Child | 452 (12.8) | 351 (40.3) | 101 (3.8) | 341 (85.3) | 111 (3.5) |
| Sibling | 92 (2.6) | 28 (3.2) | 64 (2.4) | 7 (1.8) | 85 (2.7) |
| Extended family | 127 (3.6) | 57 (6.6) | 70 (2.6) | 39 (9.8) | 88 (2.8) |
| Friend | 269 (7.6) | 22 (2.5) | 247 (9.2) | 2 (0.5) | 267 (8.5) |
| Roommate | 54 (1.5) | 1 (0.1) | 53 (2.0) | 0 (0.0) | 54 (1.7) |
| Coworker | 214 (6.0) | 29 (3.3) | 185 (6.9) | 0 (0.0) | 214 (6.8) |
| Unknown/Other | 1864 (52.5) | 274 (31.5) | 1565 (58.5) | 11 (2.8) | 1853 (58.9) |
| Age of SOI |  |  |  |  |  |
| <18 | 400 (11.3) | 364 (41.8) | 36 (1.4) | 400 (100) | 0 (0.0) |
| 18-29 | 242 (6.8) | 51 (5.9) | 191 (7.1) | 0 (0.0) | 242 (7.7) |
| 30-39 | 245 (6.9) | 58 (6.7) | 187 (7.0) | 0 (0.0) | 245 (7.8) |
| 40-49 | 191 (5.4) | 45 (5.2) | 146 (5.5) | 0 (0.0) | 191 (6.1) |
| 50-59 | 185 (5.2) | 39 (4.5) | 146 (5.5) | 0 (0.0) | 185 (5.9) |
| 60-69 | 79 (2.2) | 12 (1.4) | 67 (1.4) | 0 (0.0) | 79 (2.5) |
| ≥70 | 36 (1.0) | 6 (0.7) | 30 (1.1) | 0 (0.0) | 36 (1.1) |
| Unknown | 2167 (61.1) | 295 (33.9) | 1872 (70.0) | 0 (0.0) | 2167 (68.9) |
| Test program |  |  |  |  |  |
| Car line/for cause | 992 (28.0) | 232 (26.7) | 760 (28.4) | 45 (11.3) | 627 (19.9) |
| Asymptomatic | 672 (19.0) | 137 (15.8) | 535 (20.0) | 45 (11.3) | 890 (28.3) |
| Community | 1881 (53.0) | 501 (57.6) | 1380 (51.6) | 253 (63.3) | 1628 (51.8) |
| Ever symptomatic? |  |  |  |  |  |
| No | 271 (7.6) | 56 (6.4) | 215 (8.0) | 25 (6.3) | 246 (7.8) |
| Yes | 3274 (92.4) | 814 (93.6) | 2460 (82.0) | 375 (93.7) | 2899 (92.2) |
| Recent travel history |  |  |  |  |  |
| In-state | 46 (1.3) | 10 (1.2) | 36 (1.4) | 4 (1.0) | 42 (1.3) |
| Out-of-state | 664 (18.7) | 149 (17.1) | 515 (19.3) | 56 (14.0) | 608 (19.3) |
| International | 107 (2.0) | 26 (3.0) | 81 (3.0) | 8 (2.0) | 99 (3.1) |
| None | 2728 (77.0) | 685 (78.7) | 2043 (76.4) | 332 (83.0) | 2396 (76.2) |
| IC vaccinated? |  |  |  |  |  |
| Yes, UTD | 1063 (30.0) | 314 (36.1) | 749 (28.0) | 166 (41.5) | 897 (28.5) |
| Yes, partial | 32 (1.0) | 9 (1.0) | 23 (0.9) | 3 (0.8) | 29 (0.9) |
| Yes, not UTD | 997 (28.1) | 258 (29.7) | 739 (27.6) | 141 (35.3) | 856 (27.2) |
| Not vaccinated | 1453 (41.0) | 289 (33.2) | 1164 (43.5) | 90 (22.5) | 1363 (43.3) |
| Vaccine type |  |  |  |  |  |
| J&J | 68 (1.9) | 20 (2.3) | 48 (1.8) | 10 (2.5) | 58 (1.8) |
| Moderna | 946 (26.7) | 264 (30.3) | 682 (25.5) | 143 (35.8) | 803 (25.5) |
| Pfizer | 1071 (30.2) | 296 (34.0) | 775 (29.0) | 155 (38.8) | 916 (29.1) |

Variables with significance against CIR or having a child SOI in either adjusted or unadjusted models are included in Supplementary Table 4. The ORs in the adjusted models are presented in Figure 3. Gender was significantly associated with a child SOI (female OR 1.7, p-value <0.0001), but not with CIR (female OR 1.1, p-value 0.3139). Workers ages 30-59 were more likely than other age groups to have a CIR (OR 2.5-5.1, p-values <0.0001), and workers aged 30-49 years were significantly more likely than other age groups to report a child as their likely SOI (OR 2.2-3.0, p-values <0.05). Asymptomatic versus “for-cause” testing was not significantly associated with CIR cases, but child SOI cases were half as likely to test via asymptomatic testing than non-child SOI cases (OR 0.5, p-value 0.0001). The variables obesity, telework status, and vaccine type were negligible following model adjustment for both outcomes of interest. Finally, the odds of a child being assigned the likely SOI increased 3.4 times for each additional child reported living in the household (p-value <0.0001). All models were highly significant (p-values <0.0001).

**Figure 3.**
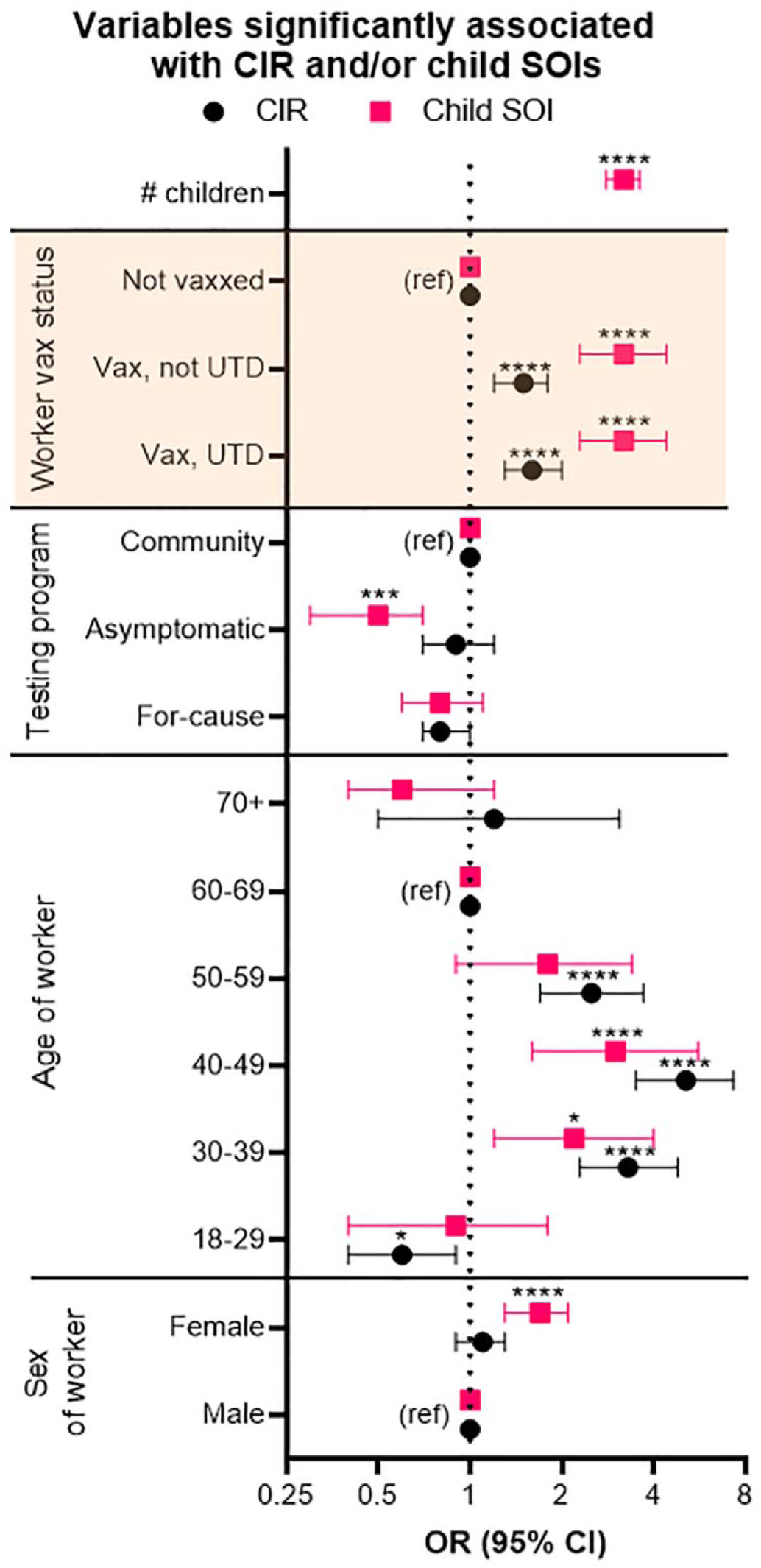
The age, sex, and vaccination status of the worker, the testing program utilized, and the number of children in the household were significantly associated with having a CIR and/or having a child SOI. Forest plot of variables found to be significantly associated with either outcome of interest following model adjustment for each variable. The outcomes of interest were either having a child in-residence (CIR) or identifying a child as the likely source of the worker’s infection (SOI). Odds ratios (ORs) were calculated based on a refernce group in each stratum (ref). Odds ratios are represented with 95% confidence intervals (CIs), and p-value significance are denoted above using the following criteria: <0.5*, <0.01**, <0.001***, <0.0001****. Data highlighted in yellow are significantly influenced by time. Raw data from both adjusted and unadjusted models are reported in Supplementary Table 4.

### Detection of variants of concern

In January 2021, a sequencing program was launched at the NIH to identify circulating VOCs in the workforce by sequencing positive samples collected by OMS. A subset of samples predating the start of this program were saved and used to characterize early circulating strains. Some samples were unable to be amplified with published primer sets (overall success generating amplicons 77.4%, Figure 4A). Sequencing following amplicon generation was 63.0% successful, and 37.0% of samples had at least 85% read alignment and were published to GISAID and the NCBI genome database.

**Figure 4.**
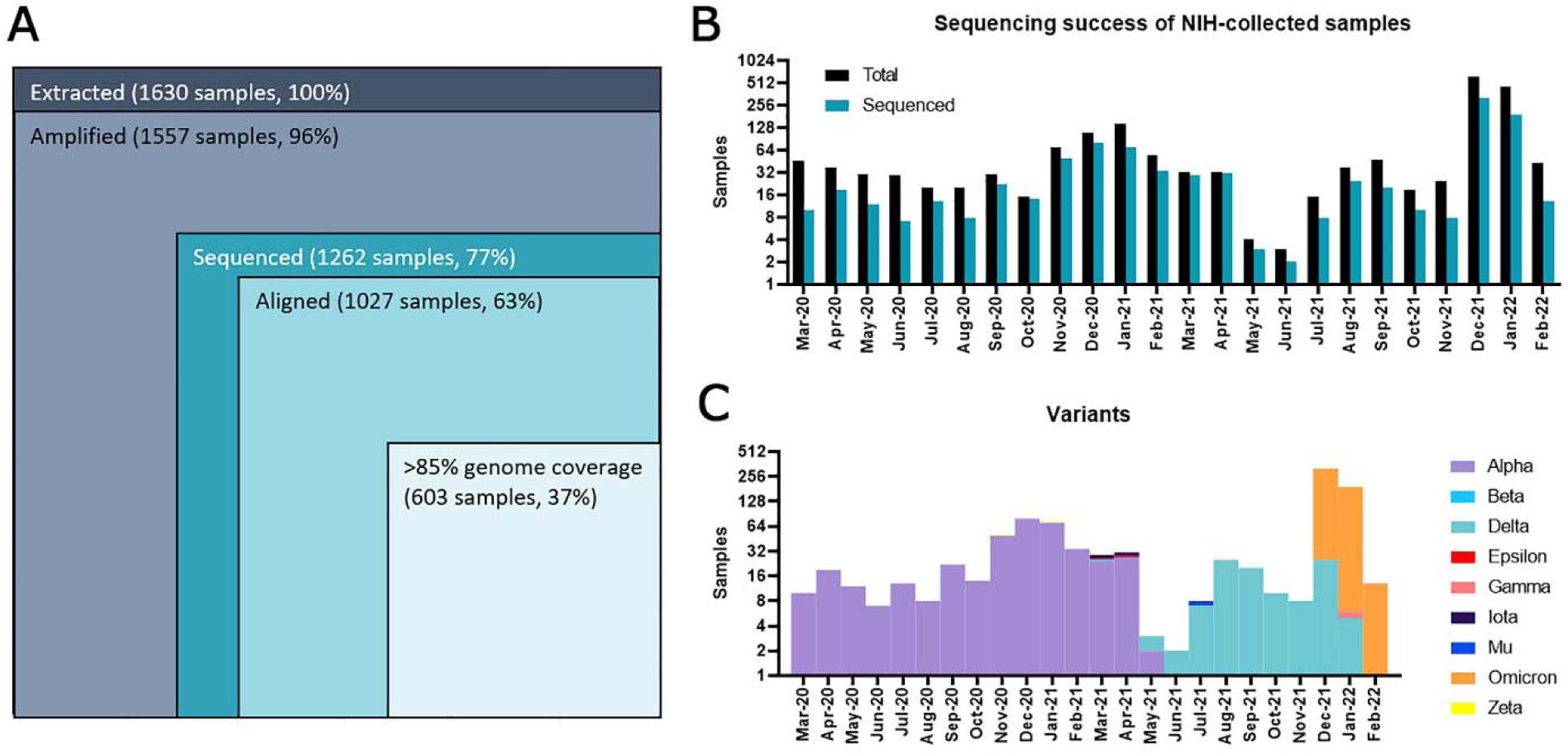
Samples collected through NIH-based testing programs were sequenced and displayed dominant VOC emergences and displacements within the study population. A) Diagram of sequencing procedure and success of each step in counts and percentages of the total NIH-collected sample set. Sample collection and sequencing (B) and VOC counts of the successfully sequenced samples (C) over time. VOC assignment followed WHO naming guidelines.

The subset of successfully sequenced samples allowing for VOC assignment was plotted by month of positive test in Figure 4B and 4C. Due to the limited number of 2020 samples that could be sequenced, the dominant VOC assignment for cases in March to December 2020 was supplemented by community-published sequences, all of which were the ancestral/Alpha variant. Delta was first detected within the study population in May 2021 and became the dominant VOC in June 2021. Omicron emerged and displaced Delta in November 2021, but low levels of Delta were detected throughout the holiday surge of 2021-2022. A drop in VOC characterization in December 2021 and January 2022 occurred for the following reasons: (1) PCR-based testing couldn’t meet increased testing demands, (2) sequencing efforts became deprioritized during surges, and (3) use of antigen-based testing increased. Therefore, dominant VOCs are aggregated monthly for analyses.

### Regression analyses

The significance of VOC impact on CIR cases compared to child SOI cases was determined using logistic regression models (Supplementary Table 5) and adjusted for age, gender, and testing program of case (Figure 5A). Child SOI cases were additionally adjusted for number of CIR. Because vaccination status and VOC are both time-dependent, vaccination status was excluded from the models. All models were highly significant (p-values <0.0001).

**Figure 5.**
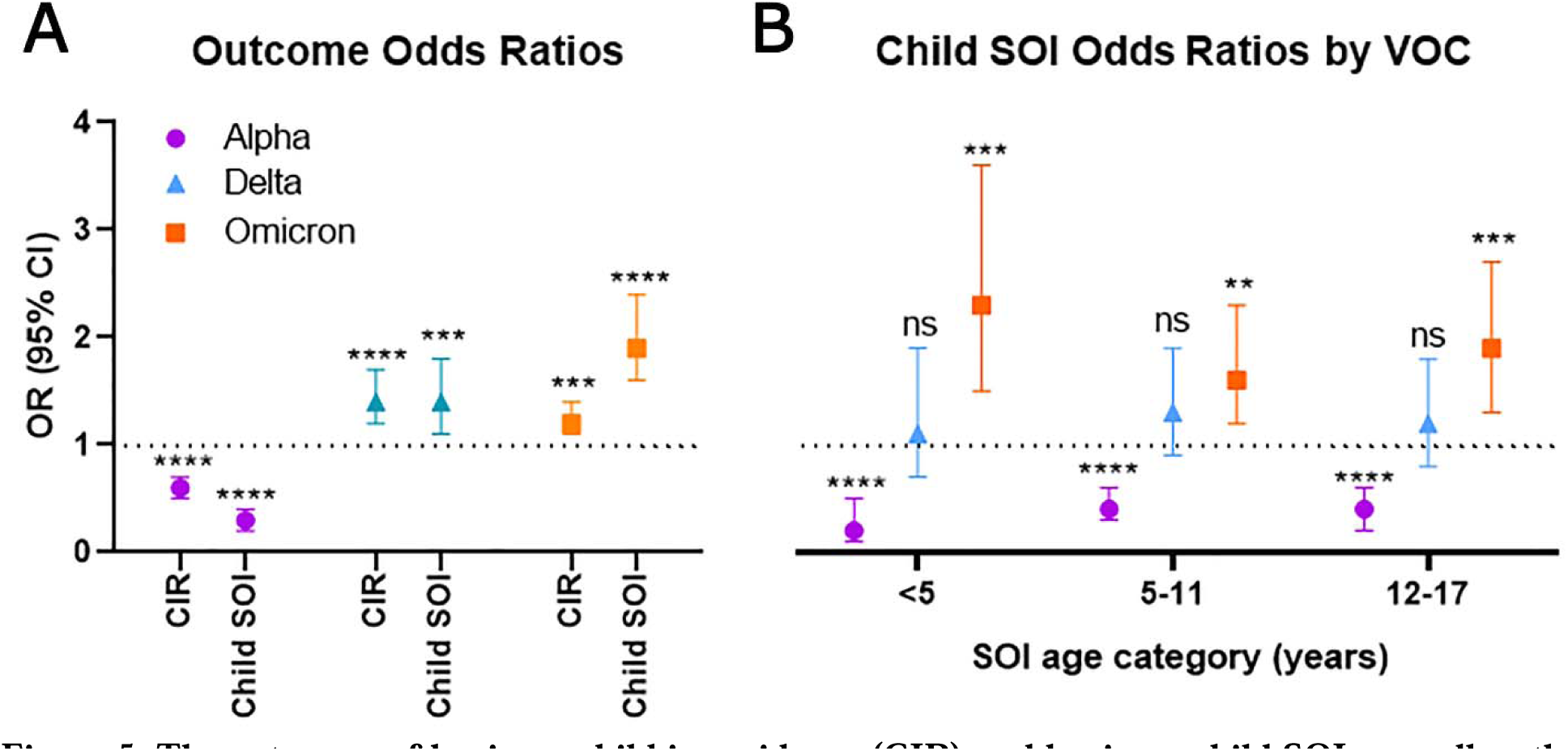
The outcomes of having a child in-residence (CIR) and having a child SOI, as well as the age of the child SOI, varied significantly with dominant VOC changes. A) Forest plot of VOC impact on odds ratios (ORs) of outcomes of interest following model adjustment for worker age, worker gender, and number of children in the household. B) Forest plot of ORs of child SOI age groups stratified by VOC, following model adjustment for worker age, worker gender, and number of children in the household. Odds ratios are represented with 95% confidence intervals (CIs), and p-value significance are denoted above using the following criteria: <0.5*, <0.01**, <0.001***, <0.0001****, ns-not significant. Raw data from both adjusted and unadjusted models are reported in Supplementary Table 5.

All dominant VOCs were significantly associated with having a CIR and having a child SOI (p-values <0.05). In Alpha-dominant months, the OR for having a CIR was 0.6 times the odds of not having a CIR, significantly lower than the odds associated with Delta- and Omicron-dominant months (ORs 1.5 and 1.2, respectively). In Alpha-dominant months, the odds of having a child SOI were 0.3 times the odds of not having a child SOI, significantly lower than the odds associated with Delta- and Omicron-dominant months (ORs 1.3 and 2.2, respectively). Thus, the odds of identifying a child SOI were half the odds of having a CIR during Alpha-dominant months, the odds of CIR and child SOIs were identical during Delta-dominant months, and the likelihood of having a child SOI was higher than having a CIR in Omicron-dominant months.

Because different child age groups became eligible for vaccination at different times, the impact of dominant VOC and the age of child SOI was explored. Children were stratified into three groups – age <5 years, 5-11 years, and 12 to 17 years - and regression models with the same adjustments previously outlined were applied to the data (Figure 5B). The odds of children 5 to 17 years old being a likely SOI were twice as high as children less than 5 years old during Alpha-dominant months (ORs 0.4 and 0.2, respectively, p-values <0.0001). The odds between each age group during Delta-dominant months were not significantly different from each other (ORs 1.1-1.3, p-values not significant). During Omicron-dominant months, odds of children under 5 being identified as the SOI were 1.4 times as high as children 5 to 11 years of age (ORs 2.4 and 1.6, respectively, p-values <0.0001).

## Discussion

We found that having a child in the household represented an important source of infection for workers during the Delta and Omicron surges as compared to the Alpha wave. We highlighted the significant burden of child SOI cases associated with female workers despite more male workers reporting a child residing at home (CIR). The SARS-CoV-2 pandemic worsened the gender gap in responsibilities related to household management and childcare, as indicated by loss of women in the workforce and their delay to return to work due to childcare needs (18). Higher odds of female workers having a likely child SOI might be explained by continued female-dominant childcare activities.

Alpha-dominant months of the COVID-19 pandemic severely impacted adults over 65 years of age (1, 4). This is consistent with our finding of low CIR during this time, as older workers were less likely to have a child below the age of 18 at home. Delta and Omicron did not appear to have the same age discrimination as their Alpha predecessor (2, 10, 19, 20), as younger workers began to be a significantly infected proportion of cases starting in mid-2021 (Supplementary Table 2). Therefore, the odds of a worker having a CIR increased during the Delta- and Omicron-dominant months.

The low child SOI odds in the early, Alpha-dominant months were likely impacted by several factors. First, research suggests that the Alpha variant may have been less virulent for children than the subsequent VOCs, resulting in more asymptomatic infections in children (1, 2, 20, 21). Second, because children were at lower risk of serious infection, testing was not as available to them early in the pandemic. Finally, children were much less likely to be exposed to the virus in the Alpha-dominant months because typical in-person activities (e.g., school, sports) were suspended until mid-2021, when Delta became the dominant VOC.

The rise in odds of having a child SOI correlated with emergence of the Delta variant, but increased travel and removal of strict mask mandates likely contributed to this trend. Loosening restrictions increased the risk of exposure among children relative to the risks during the Alpha-dominant months (22). Resumption of in-person schooling for children coincided with increased child-centric surveillance testing, which resulted in early and increased detection of prodromal and asymptomatic cases (23, 24). Increased testing likely amplified household awareness of infections. This social change would also explain why we estimated that school-aged children ages 5 to 17 comprised a higher burden of child SOI cases compared to children age <5 during that time.

Personal risk assessments during Omicron-dominant months likely influenced testing patterns, as at-home antigen testing became more available and popular. Negative results from these less sensitive antigen tests could skew SOI assignments (25, 26), especially for children that were difficult to sample, asymptomatic, or had low viral loads. Nevertheless, the risk the Omicron VOC posed to children and their adult caretakers is strongly reflected in these data. As Omicron continues to be the dominant VOC, risk posed to children and adult caretakers remain high. Future studies of surges can assess this ongoing risk.

Limitations of our interpretations are primarily due to self-report mechanisms and population of this study. Total case counts were likely impacted as some positive results were likely not reported to OMS, and workers may not have sought COVID-19 testing for vague, mild, or quickly resolving symptoms. Self-report also likely impacted child SOI assignment likelihood, as workers may have believed their children could not transmit SARS-CoV-2 and failed to attribute the SOI to them, or they may not have noticed their children were symptomatic due to decreased awareness of their risk. Overestimation of child SOIs may have taken place if parents were more likely to attribute the SOI to a child than admit to risky behavior (e.g., traveling or attending a large event). Additionally, children with minor or no recognizable symptoms may not have enough viral load to be recognized by some testing mechanisms (27, 28) and thus may have brought the virus into the household undetected. Fewer CIR and child SOI cases would ultimately mean our results were based on underestimates of both conditions.

Remaining limitations were identified as missing data from both the study population and the sequencing data. Because SOI details were not explicitly asked on the survey, workers who were unresponsive during contact tracing could not provide SOI descriptions and were instead assigned a “community” SOI. Therefore, some “community” SOIs may instead be child SOIs, leading to an overall underestimation of this outcome. Additionally, about half of the cohort reported their first positive COVID-19 test was conducted outside of the NIH. Therefore, there was no biological sample available for sequencing. Thus, dominant VOCs over time were estimated using samples from less than half of the study population and could not be attributed directly to child SOI cases.

Despite these limitations, a cohort study of this size was essential to evaluate early assertions concerning COVID-19 risks both to and from children. This study determined that children were significant SOIs to workers as Delta and Omicron became dominant, which is important for risk assessment of all child caretakers. These data support the argument for prudent use of testing, vaccination, and masking precautions for children during community surges, as the findings underscore the impact of their infections on a workforce. Further, they inform human resource policy conducive to occupational risk management to prevent secondary workplace transmission. Overall, these findings serve as an updated assessment of the significant role of children in the transmission of SARS-CoV-2, which can be extended to other common airborne pathogens liable to impact workforces.

## Human subjects

No human subjects were directly involved in this research. Consent of the participants was implied by return of the questionnaires and submission of biological samples for testing and sequencing as outlined in the Privacy Act Notice. All subjects were over the age of 18 at the time of sample and data collection.

## Author contributions

JMvLS and JLK conceived the paper and analyzed the data. HBB and MRB aided in the acquisition and interpretation of the data. JME created and edited the privacy act notice, as well as provided funding for the COVID-19 case management system which supported the data and case result data. SD led the diagnostic testing and processing of the NIH-collected specimens. AER and EG led the sequencing and genomic analysis of the specimens. MM led the data capture and integrity team. JMvLS, JLK, HBB, MRB, ADC, and DRP contributed to writing and finalizing the manuscript. All authors contributed to the editing of the manuscript.

## Data Availability

All data produced in the present study are available upon reasonable request to the authors

## Acknowledgements

We would like to thank the tireless efforts of all the teams involved in the acquisition of data and samples, especially the workers at DLM/DTM and in Elodie Ghedin’s lab for sequencing, and the OMS contact tracers including volunteers from Clinical Center Nursing Department and other NIH research and clinical staff, led by Meghan Murray, Chris Gagnon, Keith Baptiste, and Kathleen Morton. We thank those involved in the capture of health data and testing logistics at HIMD, especially Amanda Grove, Josanne Revoir, Tricia Coffey, and Jon McKeeby. Finally, we would like to thank those responsible for the data capture and integrity, especially Samantha Hughes, Courtney Bell and the OIIT team led by Judy Chan QA/QI/RM. The authors thank Yolanda L. Jones, NIH Library, for manuscript editing assistance.

---

**Supplementary Table 1.**
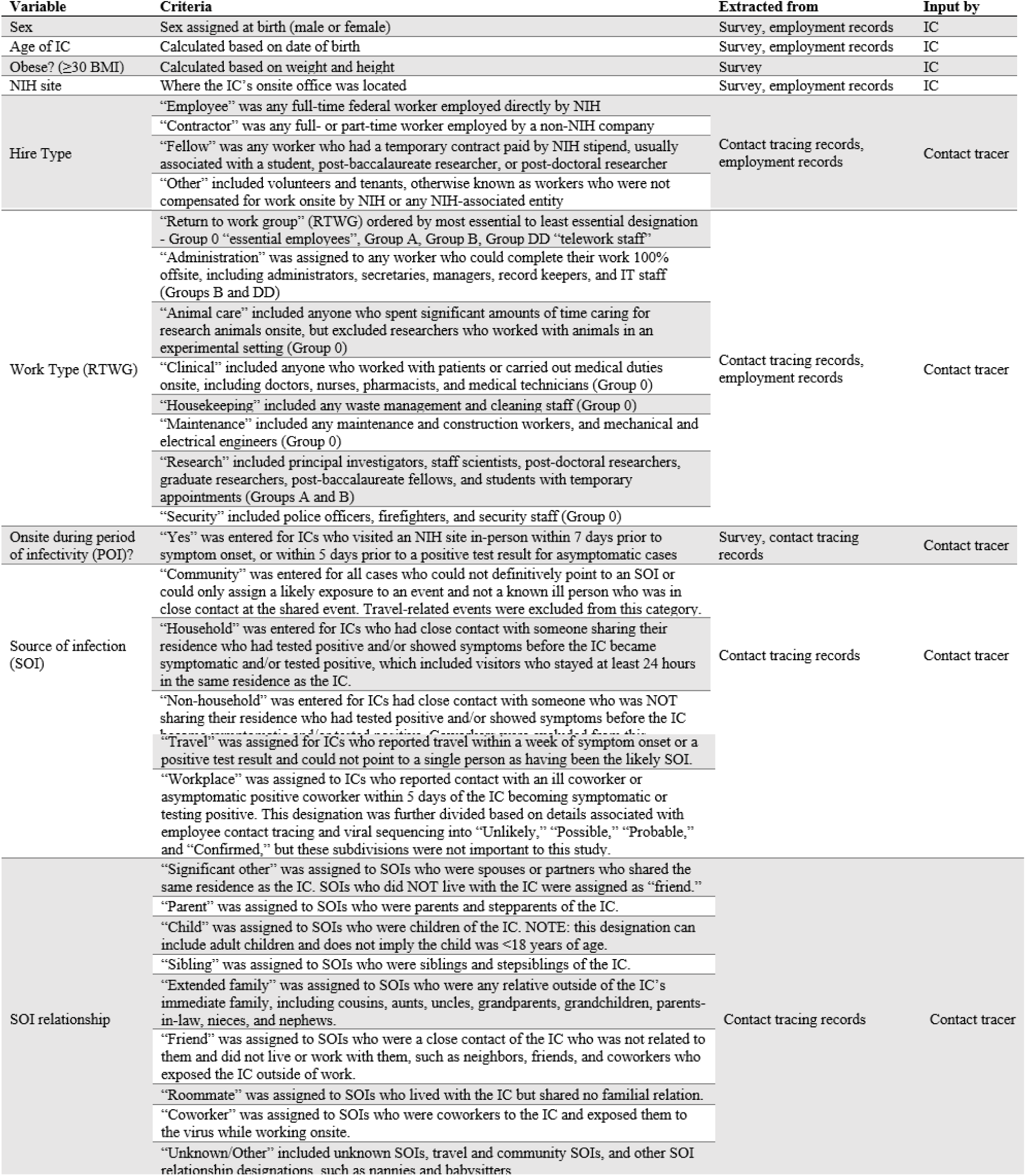
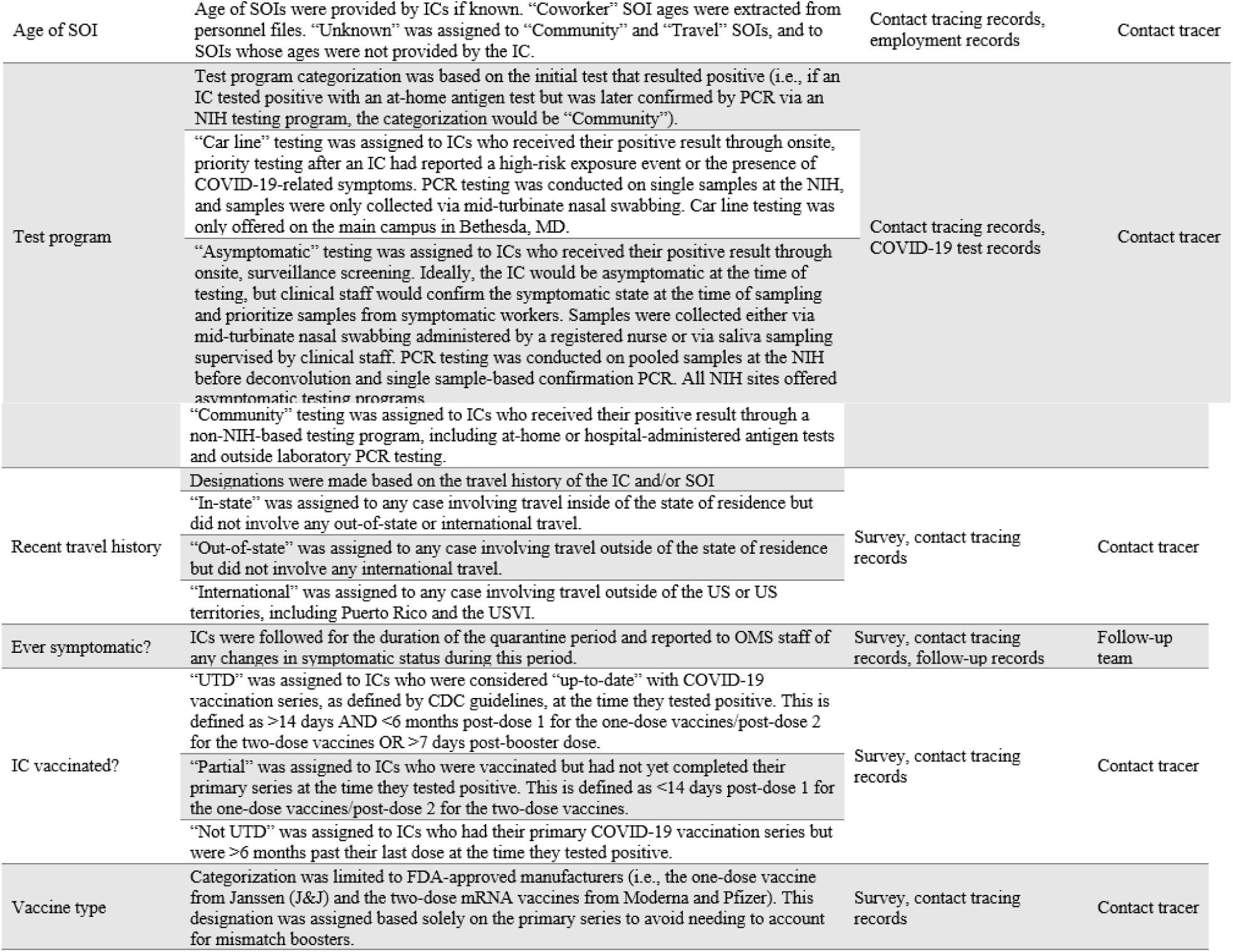
Variable categories, criteria, and extraction sources. The source(s) of data are included both in electronic records and who input the data into the records.

**Supplementary Table 2.** Descriptive statistics for the study population, unstratified and stratified by time group (TGrp). Counts and percentages are reported for each stratum.

|  | Cumulative<br>TGrp | 2020 |  |  |  | 2021 |  |  |  | 2022 |  |
| --- | --- | --- | --- | --- | --- | --- | --- | --- | --- | --- | --- |
| Variable |  | 1 | 2 | 3 | 4 | 5 | 6 | 7 | 8 | 9 | 10 |
| Total cases | 3544 (100) | 185 (5.2) | 130 (3.7) | 115 (3.2) | 423 (11.9) | 298 (8.4) | 157 (4.4) | 125 (3.5) | 151 (4.2) | 884 (24.9) | 1077 (30.4) |
| Sex |  |  |  |  |  |  |  |  |  |  |  |
| Male | 1722 (48.6) | 90 (48.7) | 68 (52.3) | 74 (64.6) | 229 (54.1) | 160 (53.7) | 69 (44.0) | 61 (48.8) | 76 (50.3) | 379 (42.9) | 516 (47.9) |
| Female | 1823 (51.4) | 95 (51.3) | 62 (47.8) | 41 (35.7) | 194 (45.9) | 138 (46.3) | 88 (56.0) | 64 (51.2) | 75 (49.7) | 505 (57.1) | 561 (52.1) |
| Age of IC |  |  |  |  |  |  |  |  |  |  |  |
| 18-29 | 710 (20.0) | 24 (13.0) | 25 (19.2) | 25 (21.7) | 79 (18.7) | 58 (19.5) | 28 (17.8) | 28 (22.4) | 21 (13.9) | 206 (23.3) | 216 (20.1) |
| 30-39 | 1015 (28.6) | 38 (20.5) | 33 (25.4) | 33 (28.7) | 113 (26.7) | 81 (27.2) | 47 (29.9) | 36 (28.8) | 42 (27.8) | 275 (31.1) | 317 (29.4) |
| 40-49 | 761 (21.5) | 51 (27.6) | 33 (25.4) | 19 (16.5) | 80 (18.9) | 61 (20.5) | 33 (21.0) | 24 (19.2) | 37 (24.5) | 182 (20.6) | 241 (22.4) |
| 50-59 | 648 (18.3) | 48 (26.0) | 24 (18.5) | 27 (23.5) | 81 (19.2) | 54 (18.1) | 28 (17.8) | 18 (14.4) | 23 (15.2) | 156 (17.7) | 189 (17.6) |
| 60-69 | 346 (10.0) | 17 (9.2) | 12 (9.2) | 9 (7.8) | 57 (13.5) | 41 (13.8) | 19 (12.1) | 14 (11.2) | 21 (13.9) | 58 (6.6) | 98 (9.1) |
| ≥70 | 52 (1.5) | 4 (2.2) | 2 (1.5) | 2 (1.7) | 11 (2.6) | 2 (0.7) | 2 (1.3) | 3 (2.4) | 7 (4.6) | 6 (0.7) | 13 (1.2) |
| Obese? (≥30 BMI) |  |  |  |  |  |  |  |  |  |  |  |
| Yes | 710 (20.0) | 50 (27.0) | 45 (34.6) | 24 (20.9) | 98 (23.2) | 70 (23.5) | 36 (22.9) | 15 (12.0) | 40 (26.5) | 159 (18.0) | 173 (16.1) |
| No | 1646 (46.4) | 68 (36.8) | 66 (50.8) | 58 (50.4) | 202 (47.8) | 122 (40.9) | 64 (40.8) | 63 (50.4) | 59 (39.1) | 412 (46.6) | 532 (49.4) |
| No info | 1189 (33.5) | 67 (36.2) | 19 (14.6) | 33 (28.7) | 123 (29.1) | 106 (35.6) | 57 (36.3) | 47 (37.6) | 52 (34.4) | 313 (35.4) | 372 (34.5) |
| NIH site |  |  |  |  |  |  |  |  |  |  |  |
| Main campus (MD) | 2458 (69.3) | 127 (68.7) | 88 (67.7) | 77 (67.0) | 243 (57.5) | 185 (62.1) | 90 (57.3) | 84 (67.2) | 95 (62.9) | 661 (74.8) | 808 (75.0) |
| Satellite sites (MD) | 825 (23.3) | 57 (30.8) | 35 (26.9) | 29 (25.2) | 137 (32.4) | 91 (30.5) | 55 (35.0) | 32 (25.6) | 36 (23.8) | 196 (22.1) | 157 (14.6) |
| NIEHS (NC) | 138 (3.9) | 0 (0.0) | 4 (3.1) | 0 (0.0) | 14 (3.3) | 16 (5.4) | 7 (4.5) | 7 (5.6) | 4 (2.7) | 19 (2.2) | 67 (6.2) |
| RML (MT) | 106 (3.0) | 0 (0.0) | 1 (0.8) | 7 (6.1) | 23 (5.4) | 6 (2.0) | 5 (3.2) | 1 (0.8) | 15 (9.9) | 6 (0.7) | 42 (3.9) |
| NIDDK (AZ) | 18 (0.5) | 1 (0.5) | 2 (1.5) | 2 (1.7) | 6 (1.4) | 0 (0.0) | 0 (0.0) | 1 (0.8) | 1 (0.7) | 2 (0.2) | 3 (0.3) |
| Hire Type |  |  |  |  |  |  |  |  |  |  |  |
| Fellow | 571 (16.1) | 7 (3.8) | 11 (8.5) | 14 (12.2) | 51 (12.1) | 44 (14.8) | 21 (13.4) | 15 (12.0) | 17 (2.0) | 154 (17.4) | 237 (22.0) |
| Employee | 1477 (41.7) | 70 (37.8) | 43 (33.1) | 32 (27.8) | 173 (40.9) | 110 (36.9) | 73 (46.5) | 60 (48.0) | 67 (44.4) | 381 (43.1) | 468 (43.5) |
| Contractor | 1415 (39.9) | 102 (55.1) | 73 (56.2) | 65 (56.5) | 183 (43.3) | 134 (45.0) | 63 (40.1) | 47 (37.6) | 64 (42.4) | 332 (37.6) | 352 (32.7) |
| Other | 82 (2.3) | 6 (3.2) | 3 (2.3) | 4 (3.5) | 16 (3.8) | 10 (3.4) | 0 (0.0) | 3 (2.4) | 3 (2.0) | 17 (1.9) | 20 (1.9) |
| Work Type (RTWG) |  |  |  |  |  |  |  |  |  |  |  |
| Administration (C) | 1043 (29.4) | 70 (37.8) | 57 (43.9) | 32 (27.8) | 159 (37.6) | 105 (35.2) | 68 (43.3) | 48 (38.4) | 42 (27.8) | 258 (29.2) | 204 (18.9) |
| Animal care (O) | 295 (8.3) | 24 (13.0) | 8 (6.2) | 15 (13.0) | 21 (5.0) | 31 (10.4) | 11 (7.0) | 10 (8.0) | 11 (7.3) | 76 (8.6) | 88 (8.2) |
| Clinical (O) | 441 (12.4) | 6 (3.2) | 7 (5.4) | 6 (5.2) | 23 (5.4) | 16 (5.4) | 12 (7.6) | 15 (12.0) | 22 (14.6) | 148 (16.7) | 186 (17.3) |
| Housekeeping (O) | 102 (2.9) | 16 (8.7) | 3 (2.3) | 7 (6.1) | 13 (3.1) | 9 (3.0) | 3 (1.9) | 1 (0.8) | 3 (2.0) | 17 (1.9) | 30 (2.8) |
| Maintenance (O) | 356 (10.0) | 21 (11.4) | 11 (8.5) | 17 (14.8) | 71 (16.8) | 35 (11.7) | 21 (13.4) | 13 (10.4) | 19 (12.6) | 55 (6.2) | 93 (8.6) |
| Research (A/B) | 1084 (30.6) | 32 (17.3) | 28 (21.5) | 25 (21.7) | 109 (25.8) | 78 (26.2) | 34 (21.7) | 33 (26.4) | 38 (25.2) | 279 (31.6) | 428 (39.7) |
| Security (O) | 156 (4.4) | 13 (7.0) | 7 (5.4) | 5 (4.4) | 21 (5.0) | 13 (4.4) | 8 (5.1) | 4 (3.2) | 13 (8.6) | 36 (4.1) | 36 (3.3) |
| Onsite during POI? |  |  |  |  |  |  |  |  |  |  |  |
| Yes | 2541 (71.7) | 122 (66.0) | 71 (54.6) | 90 (78.3) | 273 (64.5) | 192 (64.4) | 95 (60.5) | 82 (65.6) | 112 (74.2) | 695 (78.6) | 809 (75.1) |
| No | 1004 (28.3) | 63 (34.0) | 59 (45.4) | 25 (21.7) | 150 (35.5) | 106 (35.6) | 62 (39.5) | 43 (34.4) | 39 (25.8) | 189 (21.4) | 268 (24.9) |
| Source of infection |  |  |  |  |  |  |  |  |  |  |  |
| Community | 1242 (35.0) | 85 (46.0) | 56 (43.1) | 38 (33.0) | 146 (34.5) | 122 (40.9) | 65 (41.4) | 47 (37.6) | 49 (32.5) | 312 (35.3) | 322 (29.9) |
| Household | 1313 (37.0) | 60 (32.4) | 42 (32.3) | 41 (35.7) | 175 (41.4) | 109 (36.6) | 56 (35.7) | 33 (26.4) | 64 (42.4) | 316 (35.8) | 417 (38.7) |
| Non-household | 400 (11.3) | 3 (1.6) | 9 (6.9) | 13 (11.3) | 48 (11.4) | 37 (12.4) | 21 (13.4) | 14 (11.2) | 10 (6.6) | 134 (15.2) | 111 (10.3) |
| Travel | 308 (8.7) | 6 (3.2) | 13 (10.0) | 9 (7.8) | 30 (7.1) | 22 (7.4) | 10 (6.4) | 31 (24.8) | 18 (11.9) | 52 (5.9) | 117 (10.9) |
| Workplace | 282 (8.0) | 31 (16.7) | 10 (7.7) | 14 (12.2) | 24 (5.7) | 8 (2.7) | 5 (3.2) | 0 (0.0) | 10 (6.6) | 70 (7.9) | 110 (10.2) |
| SOI relationship |  |  |  |  |  |  |  |  |  |  |  |
| Significant other | 367 (10.4) | 25 (13.5) | 16 (12.3) | 22 (19.1) | 51 (12.1) | 48 (16.1) | 21 (13.4) | 12 (9.6) | 16 (10.6) | 57 (6.5) | 99 (9.2) |
| Parent | 106 (3.0) | 13 (7.0) | 1 (0.8) | 3 (2.6) | 26 (6.15) | 14 (4.7) | 5 (3.2) | 3 (2.4) | 3 (2.0) | 16 (1.8) | 22 (2.0) |
| Child | 452 (12.8) | 6 (3.2) | 7 (5.4) | 6 (5.2) | 40 (9.5) | 18 (6.0) | 18 (11.5) | 9 (7.2) | 33 (21.9) | 140 (15.8) | 175 (16.3) |
| Sibling | 92 (2.6) | 2 (1.1) | 6 (4.6) | 3 (2.6) | 18 (4.3) | 9 (3.0) | 5 (3.2) | 2 (1.6) | 6 (4.0) | 22 (2.5) | 19 (1.8) |
| Extended family | 127 (3.6) | 1 (0.5) | 5 (3.9) | 3 (2.6) | 19 (4.5) | 14 (4.7) | 11 (7.0) | 8 (6.4) | 8 (5.3) | 32 (3.6) | 26 (2.4) |
| Friend | 269 (7.6) | 1 (0.5) | 8 (6.1) | 8 (7.0) | 38 (9.0) | 17 (5.7) | 7 (4.5) | 11 (8.8) | 3 (2.0) | 94 (10.6) | 84 (7.8) |
| Roommate | 54 (1.5) | 3 (1.6) | 3 (2.3) | 2 (1.7) | 7 (1.7) | 3 (1.0) | 5 (3.2) | 1 (0.8) | 1 (0.7) | 12 (1.4) | 17 (1.6) |
| Coworker | 214 (6.0) | 24 (13.0) | 7 (5.4) | 14 (12.2) | 17 (4.0) | 6 (2.0) | 3 (1.9) | 0 (0.0) | 8 (5.3) | 51 (5.8) | 84 (7.8) |
| Unknown | 1864 (52.5) | 110 (59.4) | 77 (59.2) | 51 (44.4) | 201 (47.5) | 166 (55.7) | 79 (50.3) | 79 (63.2) | 72 (47.7) | 448 (50.7) | 541 (50.2) |
| Age of SOI |  |  |  |  |  |  |  |  |  |  |  |
| <18 | 400 (11.3) | 3 (1.6) | 9 (6.9) | 5 (4.4) | 18 (4.3) | 16 (5.4) | 17 (10.8) | 9 (7.2) | 35 (23.2) | 125 (14.1) | 163 (15.1) |
| 18-29 | 242 (6.8) | 6 (3.2) | 8 (6.2) | 10 (8.7) | 39 (9.2) | 13 (4.4) | 16 (10.2) | 6 (4.8) | 7 (4.6) | 75 (8.5) | 62 (5.8) |
| 30-39 | 245 (6.9) | 14 (7.6) | 7 (5.4) | 11 (9.6) | 37 (8.8) | 26 (8.7) | 14 (8.9) | 9 (7.2) | 9 (6.0) | 45 (5.1) | 73 (6.8) |
| 40-49 | 191 (5.4) | 17 (9.2) | 8 (6.2) | 9 (7.8) | 34 (8.0) | 19 (6.4) | 9 (5.7) | 2 (1.6) | 8 (5.3) | 32 (3.6) | 53 (4.9) |
| 50-59 | 185 (5.2) | 19 (10.3) | 8 (6.2) | 9 (7.8) | 25 (5.9) | 23 (7.7) | 11 (7.0) | 2 (1.6) | 7 (4.6) | 27 (3.1) | 54 (5.0) |
| 60-69 | 79 (2.2) | 4 (2.2) | 2 (1.5) | 5 (4.4) | 11 (2.6) | 9 (3.0) | 3 (1.9) | 6 (4.8) | 7 (4.6) | 13 (1.5) | 19 (1.8) |
| ≥70 | 36 (1.0) | 5 (2.7) | 0 (0.0) | 2 (1.7) | 6 (1.9) | 3 (1.0) | 2 (1.3) | 1 (0.8) | 3 (2.0) | 4 (0.5) | 8 (0.7) |
| Unknown | 2167 (61.1) | 117 (63.2) | 88 (67.7) | 64 (55.7) | 251 (59.3) | 189 (63.4) | 85 (54.1) | 90 (72.0) | 75 (49.7) | 563 (63.7) | 645 (59.9) |
| Child in household? |  |  |  |  |  |  |  |  |  |  |  |
| No | 2675 (75.5) | 174 (94.1) | 110 (84.6) | 95 (82.6) | 343 (81.1) | 232 (77.9) | 109 (69.4) | 84 (67.2) | 98 (64.9) | 642 (72.6) | 788 (73.2) |
| Yes | 870 (24.5) | 11 (5.9) | 20 (15.4) | 20 (17.4) | 80 (18.9) | 66 (22.2) | 48 (30.6) | 41 (32.8) | 53 (35.1) | 242 (27.4) | 289 (26.8) |
| Ever symptomatic? |  |  |  |  |  |  |  |  |  |  |  |
| No | 271 (7.6) | 5 (2.7) | 8 (6.2) | 7 (6.1) | 19 (4.5) | 16 (5.4) | 13 (8.3) | 4 (3.2) | 7 (4.6) | 74 (8.4) | 118 (11.0) |
| Yes | 3274 (92.4) | 180 (97.3) | 122 (93.9) | 108 (93.9) | 404 (95.5) | 282 (94.6) | 144 (91.7) | 121 (96.8) | 144 (95.4) | 810 (91.6) | 959 (89.0) |
| Recent travel history |  |  |  |  |  |  |  |  |  |  |  |
| In-state | 46 (1.3) | 0 (0.0) | 4 (3.1) | 2 (1.7) | 7 (1.7) | 0 (0.0) | 3 (1.9) | 0 (0.0) | 3 (2.0) | 15 (1.8) | 11 (1.0) |
| Out-of-state | 664 (18.7) | 10 (5.4) | 15 (11.5) | 15 (13.0) | 63 (14.9) | 31 (10.4) | 21 (13.4) | 52 (41.6) | 36 (23.8) | 158 (17.9) | 263 (24.4) |
| International | 107 (2.0) | 3 (1.6) | 2 (1.5) | 3 (2.6) | 4 (1.0) | 7 (2.4) | 7 (4.4) | 12 (9.6) | 7 (4.6) | 14 (1.6) | 48 (4.5) |
| None | 2728 (77.0) | 172 (93.0) | 109 (83.9) | 95 (82.6) | 349 (82.5) | 260 (87.3) | 126 (80.3) | 61 (48.8) | 105 (69.5) | 696 (78.7) | 755 (70.1) |
| IC vaccinated? |  |  |  |  |  |  |  |  |  |  |  |
| Yes, UTD | 1063 (30.0) | 0 (0.0) | 0 (0.0) | 0 (0.0) | 0 (0.0) | 0 (0.0) | 8 (5.1) | 66 (52.8) | 47 (31.1) | 331 (37.4) | 611 (56.7) |
| Yes, partial | 32 (1.0) | 0 (0.0) | 0 (0.0) | 0 (0.0) | 0 (0.0) | 6 (2.0) | 15 (9.6) | 2 (1.6) | 1 (0.7) | 5 (0.6) | 3 (0.3) |
| Yes, not UTD | 997 (28.1) | 0 (0.0) | 0 (0.0) | 0 (0.0) | 0 (0.0) | 0 (0.0) | 10 (6.4) | 13 (10.4) | 61 (40.4) | 331 (37.4) | 420 (39) |
| Not vaccinated | 1453 (41.0) | 185 (100) | 130 (100) | 115 (100) | 423 (100) | 292 (98.0) | 124 (79.0) | 44 (35.2) | 42 (27.8) | 55 (6.2) | 43 (4.0) |
| Vaccine type |  |  |  |  |  |  |  |  |  |  |  |
| J&J | 68 (1.9) | NA | NA | NA | NA | 0 (0.0) | 0 (0.0) | 4 (3.2) | 6 (4.0) | 29 (3.3) | 29 (2.7) |
| Moderna | 946 (26.7) | NA | NA | NA | NA | 6 (2.0) | 18 (11.5) | 32 (25.6) | 46 (30.5) | 350 (39.6) | 494 (45.9) |
| Pfizer | 1071 (30.2) | NA | NA | NA | NA | 0 (0.0) | 15 (9.6) | 45 (36.0) | 57 (37.8) | 449 (50.8) | 505 (46.9) |

**Supplementary Table 3.** Descriptive statistics for the study population stratified by SOI age categories and child SOI age groups. Counts and percentages are reported for each stratum.

| Variable | Unknown | All SOI age groups |  |  |  |  |  |  | Child SOI age groups |  |  |
| --- | --- | --- | --- | --- | --- | --- | --- | --- | --- | --- | --- |
|  |  | <17 | 18-29 | 30-39 | 40-49 | 50-59 | 60-69 | >70 | <5 | 5-11 | 12-17 |
| Total cases | 2167 (61.1) | 400 (11.2) | 242 (6.8) | 245 (6.9) | 191 (5.3) | 185 (5.2) | 79 (2.2) | 36 (1.0) | 95 (2.7) | 155 (4.4) | 150 (4.2) |
| Sex |  |  |  |  |  |  |  |  |  |  |  |
| Male | 1107 (51.1) | 148 (37.0) | 113 (46.7) | 113 (46.1) | 92 (48.2) | 92 (49.7) | 41 (51.9) | 16 (44.4) | 36 (37.9) | 59 (38.1) | 53 (35.3) |
| Female | 1060 (48.9) | 252 (63.0) | 129 (53.3) | 132 (53.9) | 99 (51.8) | 93 (50.3) | 38 (48.1) | 20 (55.6) | 59 (62.1) | 96 (61.9) | 97 (64.7) |
| Age of IC |  |  |  |  |  |  |  |  |  |  |  |
| 18-29 | 496 (22.9) | 26 (6.5) | 100 (41.3) | 23 (9.4) | 47 (24.6) | 9 (4.9) | 8 (10.1) | 1 (2.8) | 8 (8.4) | 8 (5.2) | 10 (6.7) |
| 30-39 | 626 (28.9) | 138 (34.5) | 46 (19.0) | 132 (53.9) | 20 (10.5) | 42 (22.7) | 7 (8.9) | 4 (11.1) | 58 (61.1) | 67 (43.2) | 13 (8.7) |
| 40-49 | 426 (19.66) | 156 (39.0) | 15 (6.2) | 39 (15.9) | 83 (43.5) | 24 (13.0) | 13 (16.5) | 5 (13.9) | 25 (26.3) | 58 (37.4) | 73 (48.7) |
| 50-59 | 369 (17.0) | 64 (16.0) | 55 (22.7) | 22 (9.0) | 26 (13.6) | 85 (46.0) | 11 (13.9) | 16 (44.4) | 2 (2.1) | 18 (11.6) | 44 (29.3) |
| 60-69 | 198 (9.1) | 14 (3.5) | 26 (10.7) | 26 (10.6) | 12 (6.3) | 25 (13.5) | 38 (48.1) | 7 (19.4) | 0 (0.0) | 4 (2.6) | 10 (6.7) |
| ≥70 | 40 (1.9) | 2 (0.5) | 0 (0.0) | 3 (1.2) | 3 (1.6) | 0 (0.0) | 1 (1.3) | 3 (8.3) | 2 (2.1) | 0 (0.0) | 0 (0.0) |
| Work Type (RTWG) |  |  |  |  |  |  |  |  |  |  |  |
| Administration (C) | 638 (29.4) | 116 (29.0) | 63 (26.0) | 73 (29.8) | 58 (30.4) | 63 (34.1) | 18 (22.8) | 14 (38.9) | 19 (20.0) | 48 (31.0) | 49 (32.7) |
| Animal care (0) | 171 (7.9) | 23 (5.8) | 35 (14.5) | 25 (10.2) | 15 (7.9) | 19 (10.3) | 6 (7.6) | 1 (2.8) | 5 (5.3) | 10 (6.5) | 8 (5.3) |
| Clinical (0) | 234 (10.8) | 89 (22.3) | 22 (9.1) | 33 (13.5) | 20 (10.5) | 27 (14.6) | 8 (10.1) | 8 (22.2) | 23 (24.2) | 38 (24.5) | 28 (18.7) |
| Housekeeping (0) | 58 (2.7) | 7 (1.8) | 9 (3.7) | 7 (2.9) | 9 (4.7) | 7 (3.8) | 4 (5.1) | 1 (2.8) | 1 (1.1) | 1 (0.7) | 5 (3.3) |
| Maintenance (0) | 196 (9.0) | 36 (9.0) | 22 (9.1) | 27 (11.0) | 21 (11.0) | 26 (14.1) | 23 (29.1) | 5 (13.9) | 5 (5.3) | 14 (9.0) | 17 (11.3) |
| Research (A/B) | 726 (33.5) | 109 (27.3) | 80 (33.1) | 64 (26.1) | 49 (25.7) | 31 (16.7) | 18 (22.8) | 7 (19.4) | 36 (37.9) | 37 (23.9) | 36 (24.0) |
| Security (0) | 98 (4.5) | 15 (3.8) | 7 (2.9) | 10 (4.1) | 14 (7.3) | 11 (6.0) | 1 (1.3) | 0 (0.0) | 5 (5.3) | 4 (2.6) | 6 (4.0) |
| SOI relationship |  |  |  |  |  |  |  |  |  |  |  |
| Significant other | 0 (0.0) | 0 (0.0) | 51 (21.1) | 124 (50.6) | 76 (38.8) | 81 (43.8) | 32 (40.5) | 3 (8.3) | 0 (0.0) | 0 (0.0) | 0 (0.0) |
| Parent | 0 (0.0) | 0 (0.0) | 0 (0.0) | 1 (0.4) | 42 (22.0) | 32 (17.3) | 11 (13.9) | 20 (55.6) | 0 (0.0) | 0 (0.0) | 0 (0.0) |
| Child | 2 (0.1) | 341 (85.3) | 74 (30.6) | 29 (11.8) | 6 (3.1) | 0 (0.0) | 0 (0.0) | 0 (0.0) | 91 (95.8) | 126 (81.3) | 124 (82.7) |
| Sibling | 0 (0.0) | 7 (1.8) | 39 (16.1) | 19 (7.8) | 15 (7.9) | 10 (5.4) | 2 (2.5) | 0 (0.0) | 0 (0.0) | 1 (0.7) | 6 (4.0) |
| Extended family | 9 (0.4) | 39 (9.8) | 18 (7.4) | 17 (6.9) | 11 (5.8) | 16 (8.7) | 10 (12.7) | 7 (19.4) | 4 (4.2) | 20 (12.9) | 15 (10.0) |
| Friend | 260 (12.0) | 2 (0.5) | 1 (0.4) | 3 (1.2) | 1 (0.5) | 2 (1.1) | 0 (0.0) | 0 (0.0) | 0 (0.0) | 1 (0.7) | 1 (0.7) |
| Roommate | 30 (1.4) | 0 (0.0) | 16 (6.6) | 5 (2.0) | 1 (0.5) | 1 (0.5) | 1 (1.3) | 0 (0.0) | 0 (0.0) | 0 (0.0) | 0 (0.0) |
| Coworker | 22 (1.0) | 0 (0.0) | 40 (16.5) | 45 (18.4) | 37 (19.4) | 43 (23.2) | 23 (29.1) | 4 (11.1) | 0 (0.0) | 0 (0.0) | 0 (0.0) |
| Other | 1822 (84.1) | 11 (2.8) | 3 (1.2) | 2 (0.8) | 2 (1.1) | 0 (0.0) | 0 (0.0) | 2 (5.6) | 0 (0.0) | 7 (4.5) | 4 (2.7) |
| Recent travel history |  |  |  |  |  |  |  |  |  |  |  |
| In-state | 37 (1.7) | 4 (1.0) | 0 (0.0) | 0 (0.0) | 1 (0.5) | 2 (1.1) | 1 (1.3) | 1 (2.8) | 0 (0.0) | 2 (1.3) | 2 (1.3) |
| Out-of-state | 458 (21.1) | 56 (14.0) | 41 (16.9) | 27 (11.0) | 29 (15.2) | 29 (15.7) | 17 (21.5) | 7 (19.4) | 12 (12.6) | 22 (14.2) | 22 (14.7) |
| International | 77 (3.6) | 8 (2.0) | 4 (1.7) | 7 (2.9) | 2 (1.1) | 4 (2.2) | 4 (5.1) | 1 (2.8) | 6 (6.3) | 2 (1.3) | 0 (0.0) |
| None | 1595 (73.6) | 332 (83.0) | 197 (81.4) | 211 (86.1) | 159 (83.3) | 150 (81.1) | 57 (72.2) | 27 (75.0) | 77 (81.1) | 129 (83.2) | 126 (84.0) |
| IC vaccinated? |  |  |  |  |  |  |  |  |  |  |  |
| Yes, UTD | 644 (29.7) | 166 (41.5) | 69 (28.5) | 68 (27.8) | 44 (23.0) | 45 (24.3) | 20 (25.3) | 7 (19.4) | 42 (44.2) | 60 (38.7) | 64 (42.7) |
| Yes, partial | 18 (0.8) | 3 (0.8) | 2 (0.8) | 4 (1.6) | 1 (0.5) | 3 (1.6) | 1 (1.3) | 0 (0.0) | 0 (0.0) | 2 (1.3) | 1 (0.7) |
| Yes, not UTD | 625 (28.8) | 141 (35.3) | 76 (31.4) | 56 (22.9) | 41 (21.5) | 35 (18.9) | 16 (20.3) | 7 (29.4) | 36 (37.9) | 54 (34.8) | 51 (34.0) |
| Not vaccinated | 880 (40.6) | 90 (22.5) | 95 (39.3) | 117 (47.8) | 105 (55.0) | 102 (55.1) | 42 (53.2) | 22 (61.1) | 17 (17.9) | 39 (25.2) | 34 (22.7) |
| Vaccine type |  |  |  |  |  |  |  |  |  |  |  |
| J&J | 40 (1.9) | 10 (2.5) | 4 (1.7) | 6 (2.5) | 4 (2.1) | 0 (0.0) | 3 (3.8) | 1 (2.8) | 1 (1.1) | 6 (3.9) | 3 (2.0) |
| Moderna | 561 (25.9) | 143 (35.8) | 65 (26.9) | 69 (28.2) | 40 (20.9) | 47 (25.4) | 14 (17.7) | 7 (19.4) | 41 (43.2) | 43 (27.7) | 59 (39.3) |
| Pfizer | 681 (31.4) | 155 (38.8) | 78 (32.2) | 53 (21.6) | 42 (22.0) | 36 (19.5) | 20 (25.3) | 6 (16.7) | 36 (37.9) | 66 (42.6) | 53 (35.3) |

**Supplementary Table 4.**
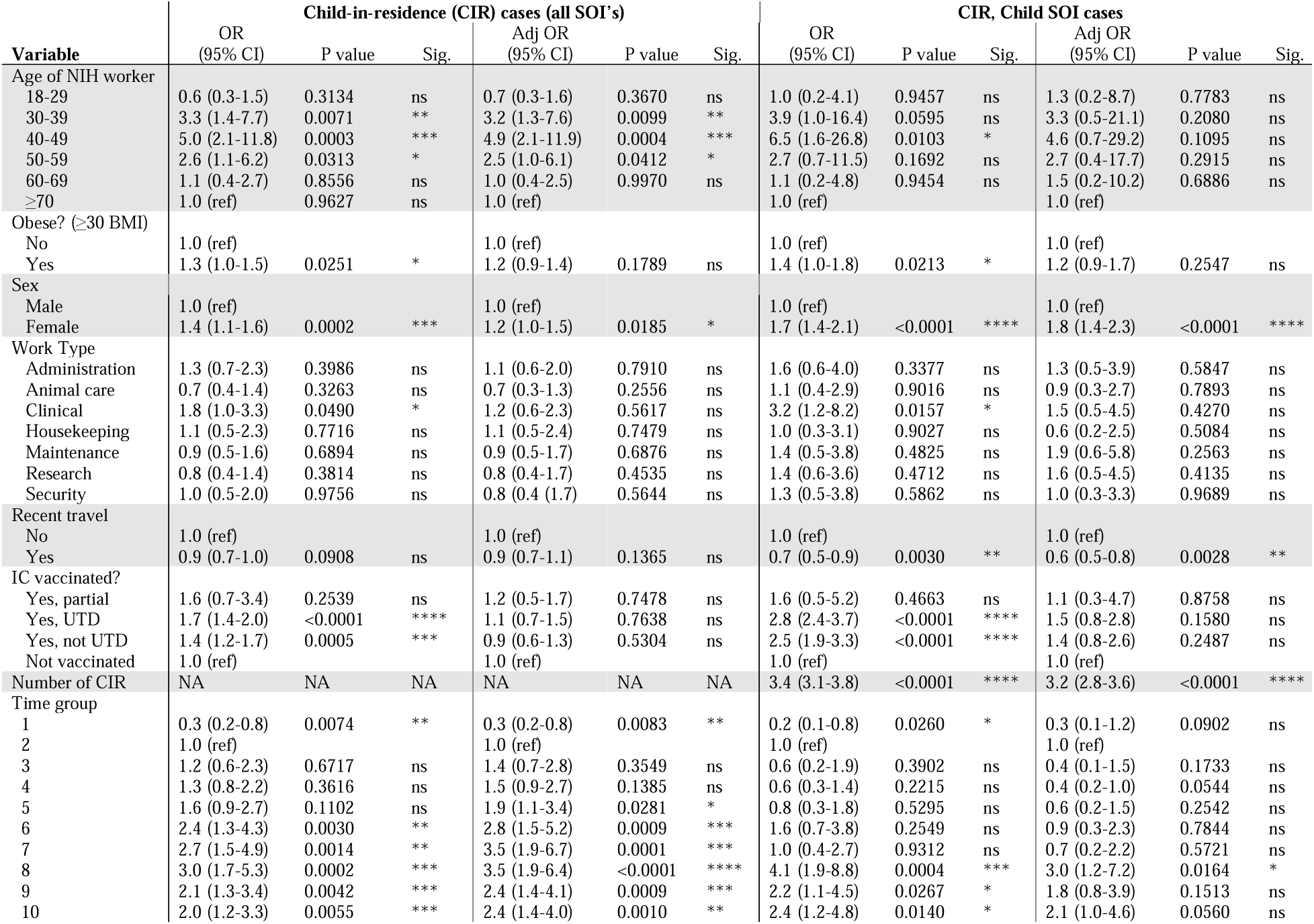
Variables significantly associated with either having a child living with a positive NIH worker (CIR) and/or having a child be the source of their infection (SOI). Odds ratios (OR) and 95% confidence intervals (CIs) were calculated in both unadjusted and adjusted (Adj) logistic regression models relative to a reference group (ref). P-values were reported for each OR and denoted for significance, p-values <0.5*, <0.01**, <0.001***, <0.0001****. Missing data were denoted with “NA.” P-values that were not significant were denoted with “ns.”

**Supplementary Table 5.** VOCs were significantly associated with either having a child living with a positive NIH worker (CIR) and/or having a child be the source of their infection (SOI). Odds ratios (OR) and 95% confidence intervals (CIs) were calculated in both unadjusted and adjusted (Adj) logistic regression models relative to a reference group (ref). P-values were reported for each OR, with significance determined to be <0.05.

| Variable | Alpha variant |  |  |  | Delta variant |  |  |  | OR<br>(95% CI) |
| --- | --- | --- | --- | --- | --- | --- | --- | --- | --- |
|  | OR<br>(95% CI) | P value | Adj OR*<br>(95% CI) | P value | OR<br>(95% CI) | P value | Adj OR*<br>(95% CI) | P value |  |
| **CIR** |  |  |  |  |  |  |  |  |  |
| No | 1.0 (ref) |  | 1.0 (ref) |  | 1.0 (ref) |  | 1.0 (ref) |  | 1.0 (ref) |
| Yes | 0.6 (0.5-0.7) | <0.0001 | 0.6 (0.5-0.7) | <0.0001 | 1.4 (1.2-1.7) | <0.0001 | 1.4 (1.2-1.7) | <0.0001 | 1.2 (1.1-1.3) |
| **Child SOI** |  |  |  |  |  |  |  |  |  |
| No | 1.0 (ref) |  | 1.0 (ref) |  | 1.0 (ref) |  | 1.0 (ref) |  | 1.0 (ref) |
| Yes | 0.3 (0.2-0.4) | <0.0001 | 0.3 (0.2-0.4) | <0.0001 | 1.4 (1.1-1.8) | 0.0026 | 1.4 (1.1-1.8) | 0.0043 | 1.9 (1.6-2.3) |
| **Age group** |  |  |  |  |  |  |  |  |  |
| <5 | 0.2 (0.1-0.4) | <0.0001 | 0.2 (0.1-0.5) | <0.0001 | 1.3 (0.8-2.1) | 0.2624 | 1.1 (0.7-1.9) | 0.6073 | 2.3 (1.5-3.5) |
| 5-11 | 0.4 (0.2-0.6) | <0.0001 | 0.4 (0.3-0.6) | <0.0001 | 1.4 (1.0-2.1) | 0.0379 | 1.3 (0.9-1.9) | 0.1954 | 1.6 (1.2-2.1) |
| 12-17 | 0.4 (0.2-0.6) | <0.0001 | 0.4 (0.2-0.6) | <0.0001 | 1.3 (1.0-2.0) | 0.0842 | 1.2 (0.8-1.8) | 0.3079 | 1.7 (1.2-2.3) |

\* Adjusted for sex and age of NIH worker.
\*\*Adjusted for sex and age of NIH worker and for the number of children in residence.

**Supplementary Figure 1.**
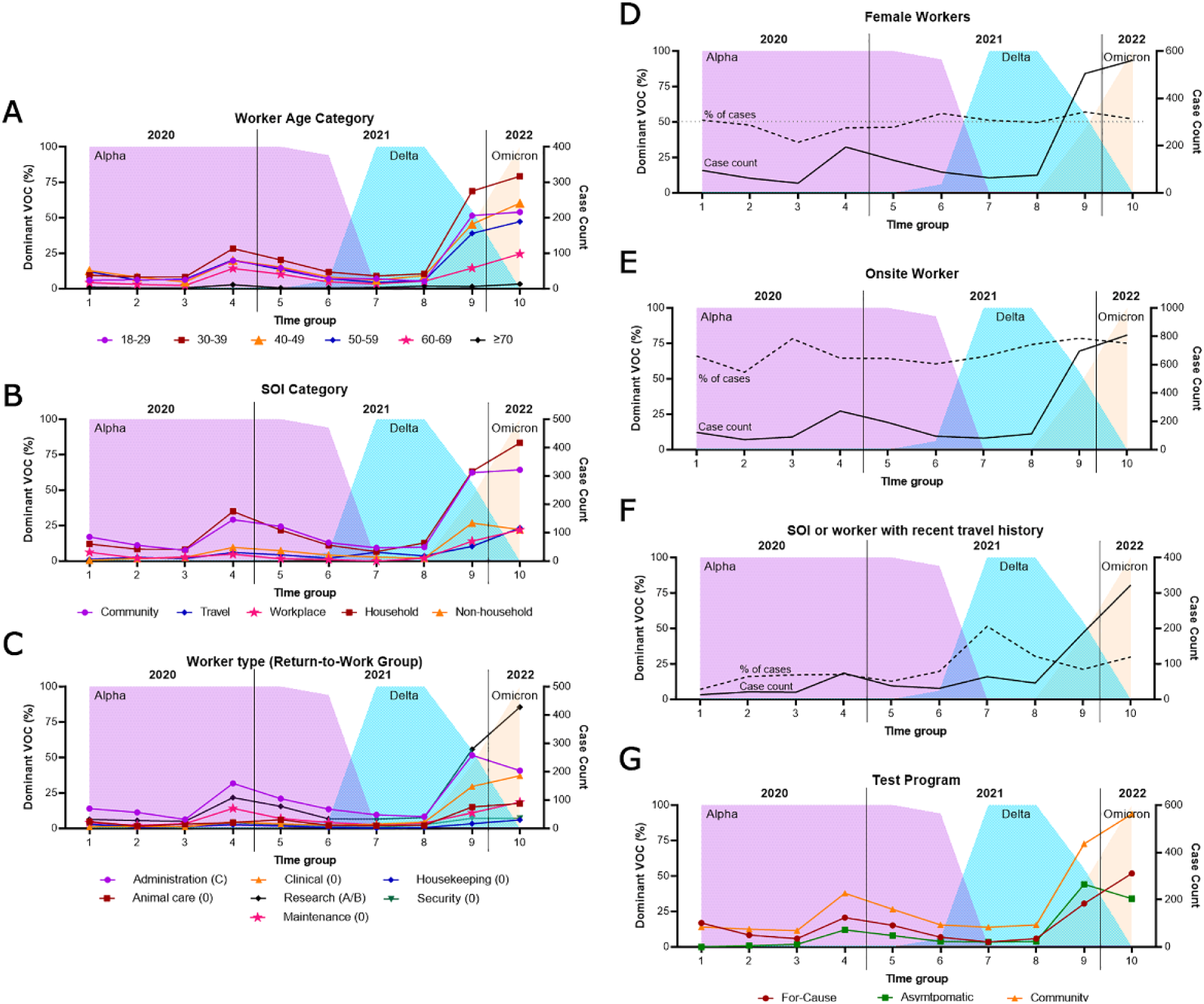
Variants of concern detected within the total NIH positive case cohort, represented in time groups and as percentages of total samples sequenced, overlayed with cumulative case counts over various strata. Strata included (A) the age category of the positive NIH worker, (B) the category of SOI assigned to the positive case, (C) the type of work conducted by the NIH worker and RTWG assignment associated with this work, (D) sex of the worker, (E) whether the worker was onsite within 7 days of symptom onset, (F) recent travel history associated with the case, and (G) testing program which gave the worker their first positive result. Raw data is reported in Supplementary Table 2.

## Abbreviations

Adj: adjusted
AZ: Arizona, USA
BMI: body mass index, = kilograms/meters^2^
CC: NIH clinical center
CDC: Centers for Disease Control and Prevention
CI: confidence interval
CIR: child(ren) in-residence
COVID-19: Coronavirus disease 2019
Ct: cycle threshold, for PCR testing
DLM: Department of Laboratory Medicine
DIR: Division of Intramural Research
FDA: Food and Drug Administration Group 0 – essential workers
Group A/B: non-essential, priority workers
Group C: teleworkers
IC: index case (NIH worker who reported a positive result)
MD: Maryland, USA
MT: Montana, USA
mRNA: messenger ribonucleic acid
NC: North Carolina, USA
NIAID: National Institutes of Allergy and Infectious Disease
NIH: National Institutes of Health
NIEHS: National Institute of Environmental Health Sciences, campus located in NC
NIDDK: National Institute of Diabetes and Digestive and Kidney Diseases, located in AZ Ns – not significant, p-value >0.05
OMS: Office of Medical Safety
OR: odds ratio
PCR: polymerase chain reaction
POI: period of infectivity
Ref: reference group, for regression analyses
RML: Rocky Mountain Labs, campus located in MT
RWTG: return-to-work group, either Group 0, Group A, Group B, or Group C
SARS-CoV- 2: severe acute respiratory syndrome coronavirus 2
Sig.: significance, p-values <0.5*, <0.01**, <0.001***, <0.0001****
SOI: source of infection
TGrp: time group
UTD: up to date, defined as >14 days post-primary series OR >7 days post-booster dose
VOC: variant of concern

